# Outlier expression of isoforms by targeted RNA sequencing as clinical markers of genomic variants in B lymphoblastic leukemia and other tumor types

**DOI:** 10.1101/2022.07.29.22278149

**Authors:** Harrison K. Tsai, Tasos Gogakos, Va Lip, Jonathan Tsai, Yen-Der Li, Adam Fisch, Jonathan Weiss, Leslie Grimmett, Thai Hoa Tran, Maxime Caron, Sylvie Langlois, Daniel Sinnett, Yana Pikman, Annette S. Kim, Valentina Nardi, Lewis B. Silverman, Marian H. Harris

## Abstract

Recognition of aberrant gene isoforms indicative of underlying DNA events can impact molecular classification and risk stratification of B lymphoblastic leukemia (B-ALL). Aberrant *ERG* isoforms have been proposed as markers of the favorable-risk *DUX4*-rearranged (*DUX4*r) subtype while deletion-mediated *IKZF1* isoforms are associated with adverse prognosis in non-*DUX4*r B-ALL. The high-risk *IKZF1*^plus^ signature depends on gene deletions including *PAX5* while intragenic *PAX5* amplifications (PAX5amp) are recurrent in the provisional B-ALL with *PAX5*-alteration subtype. In this study, we screened for outlier expression of isoforms within targeted RNA sequencing assays designed for fusions. Outlier analysis of known and novel *IKZF1*, *ERG*, and *PAX5* isoforms was 97.0% (32/33), 90% (9/10), and 100% (6/6) sensitive and 97.8% (226/231), 100% (35/35), and 88.5% (23/26) specific for *IKZF1* intragenic or 3’ deletions, *DUX4*r, and *PAX5* intragenic deletions respectively, where false positives were favored to represent low-level deletions below the limit of DNA-based detection. Outlier analysis also identified putative PAX5amp cases and revealed partial tandem duplication (PTD) spanning *IKZF1* N159Y in the B-ALL with mutated N159Y subtype. To demonstrate utility in other tumor types, outlier analysis was 100% (9/9) sensitive and 100% (255/255) specific for *KMT2A*-PTD in hematologic samples and 100% (7/7) sensitive and 100% (79/79) specific for *FGFR1* tyrosine kinase domain duplication in brain tumors. These findings support the use of aberrant isoform analysis in targeted RNA sequencing data as a robust strategy for the detection of clinically significant DNA events.

## Introduction

The versatile applications of gene fusions associated with chromosomal rearrangements towards diagnosis, prognostication, treatment, and surveillance of disease has motivated a growing use of targeted RNA sequencing in clinical laboratories. Gene isoforms, characterized by exon skipping, out-of-order exons, cryptic splice sites, or other aberrations, may similarly reflect deletions, duplications, spice variants, or other underlying DNA alterations of clinical importance, though the use of targeted RNA sequencing for purpose of their identification is less well established. Molecular classification and risk stratification of pediatric B lymphoblastic leukemia (B-ALL) may particularly benefit from recognition of aberrant isoforms. *ERG* isoforms induced by *DUX4* binding (ERGaltA/B) or mediated by intragenic *ERG* deletions have been proposed as markers of the favorable-risk *DUX4* rearranged (*DUX4*r) subtype, which occurs in 4-14% of pediatric B-ALL but has typically required whole genome sequencing or whole transcriptome RNA sequencing (wtRNAseq) for detection^1–3^. In non-*DUX4r* B-ALL, dominant negative and loss-of-function *IKZF1* isoforms arising from intragenic *IKZF1* deletions have been associated with a worse prognosis while *PAX5* isoforms with out-of-order exons have been associated with intragenic *PAX5* amplification (PAX5amp) within the emerging *PAX5*-altered subtype (PAX5alt)^4–6^. Patterns of isoform co-occurrence also carry clinical significance, highlighted by the high-risk signature *IKZF1*^plus^ defined by various co-deletions^7–9^. Several clinically important DNA variants generate aberrant RNA isoforms outside of B-ALL, including *KMT2A*-PTD in myeloid malignancies, *FGFR1* tyrosine kinase domain duplication (*FGFR1*-TKDD) in low grade gliomas, and many others^10–11^.

The isoforms described above each harbor an isoform-specific junction, however RNA transcripts from sources other than DNA aberrations can a priori contain identical junctions. Native alternative splicing and alternative transcriptional initiation are theoretically capable of producing exon-skipping isoforms and ERGalt transcripts, while circular RNAs (circRNAs) might produce the out-of-order exons of PAX5amp or *KMT2A*-PTD. Since these other sources generally produce RNA at low levels, characterization of their empiric background distribution is important to optimize sensitivity and specificity of aberrant isoforms as NGS markers of DNA events. In this study, we apply systematic outlier analysis to targeted RNA next generation sequencing (NGS) data based on anchored multiplex PCR (AMP), a popular enrichment method for detecting fusions^12^. We validate outlier expression of select isoforms as markers of underlying genomic variants and more generally describe informatic strategies for evaluating intragenic variants within a clinical practice.

## Methods

### Sample selection for targeted RNA NGS

Total nucleic acid extracted from bone marrow, peripheral blood, cell lines, or formalin-fixed paraffin-embedded tissue of other disease sites was tested at Boston Children’s Hospital by 1 of 2 clinically validated targeted NGS panels based on AMP (ArcherDx, Boulder, CO): i) ArcherDx FusionPlex Heme v2 assay (FPH) performed clinically, for research, or for validation on hematologic samples from 2014-2022 (heme cohort; n = 407) comprising predominantly pediatric cases but also including known *KMT2A*-PTD adult samples and the EOL-1 cell line or ii) Custom Solid and Brain Tumor Fusion Panel (SBT) performed clinically on pediatric brain samples (n=86) and solid tumor samples (n=95) from 2020-2021, each followed by 2 x 151 bp paired-end Illumina sequencing and bioinformatic processing by Archer Analysis v6.2.7 (AA) under default parameters. NGS samples yielding under 25,000 unique RNA reads were excluded from the above cohorts. The study was conducted in accordance with the Helsinki Declaration and with approval of Institutional Review Boards.

### Isoform junction notation, split-read analysis, and outlier criteria

We used the notation *IKZF1* e3e8 to denote the junction connecting the 3’ end of *IKZF1* exon 3 to the 5’ end of exon 8, and similarly for other genes and exact junctions. For each gene of interest, paired reads were extracted from binary alignment map files if they aligned within 1 Mb of the gene locus and involved both a primary and a supplementary alignment for at least one read of the pair (i.e. a split-read). Primary and supplementary alignments were parsed to determine the genomic coordinates (chrA:x0 and chrA:x1) indicating a breakpoint via soft-clipping or hard-clipping. In this manner, split-reads were associated with isoform junctions of the form “chrA:x0_x1_overlap_n”, where overlap was the amount of microhomology (n>0) or unaligned nucleotides (n<0) around the breakpoints. Paired reads assigned to each isoform junction were counted and sorted further by originating gene specific primer (GSP2), which was inferred from the alignment start coordinates of anchored second mate reads. Based on these paired-read assignments, expressed variant allele fraction (VAF) of an isoform junction was calculated relative to all isoform junctions sharing a breakpoint within 10 bp but restricted to split-reads originating from a GSP2 targeting the appropriate side of the shared breakpoint. Normalized expression of an isoform junction was defined by dividing split-read counts by total unique RNA reads. Isoform junctions were annotated by comparing against exon coordinates of transcripts from NCBI Refseq and Gencode to test equivalence with exact exon-exon junctions; otherwise, an isoform junction was considered novel and potentially represented use of a cryptic splice site. Isoform junctions with both breakpoints contained within a single intron of all known transcripts were predicted to have minimal effect and were filtered out. For remaining isoform junctions, outlier expression was defined as satisfying both VAF ≥ max(0.01, 5[90% quantile VAF expression]) and normalized expression ≥ 0.0001.

### Whole-transcriptome RNA-sequencing for *DUX4*r status

*DUX4*r status was available for select cases (n=45) from the heme cohort based on clinical reports from whole transcriptome RNA-sequencing tests performed at Centre Hospitalier Universitaire Sainte Justine^13^.

### DNA-based testing

Most (n=264) of the heme cohort and all pediatric brain samples underwent concurrent clinical testing by 1 of 2 DNA-based targeted NGS panels performed clinically at Brigham and Women’s Hospital: (i) Rapid Heme Panel version 3 (RHP) based on NEBNext Direct capture/amplicon hybrid chemistry (NEB; New England BioLabs, Ipswich, MA)^14^ or (ii) Oncopanel (OP) based on Agilent SureSelect hybridization capture probes (Agilent Technologies, Santa Clara, CA)^15^. RHP and OP were clinically validated to detect copy number changes of *IKZF1*, *ERG*, and *KMT2A*, and OP was also validated to detect structural rearrangements of *FGFR1.* RHP did not routinely report involved exons, thus deletion status of *IKZF1* and *ERG* on the level of individual targeted exons was re-interpreted independently by 3 molecular pathologists (HKT, MHH, TSG) based on supplementary analysis of read count data (modified BatchCNV)^16^. Copy number of *PAX5*, *IKZF1*, and *ERG* were additionally available for a limited subset (n=32) of the heme cohort based on (i) clinical reports from cytogenetic microarray testing (CGX Onco Array; PerkinElmer, Waltham, MA) performed at Centre Hospitalier Universitaire Sainte Justine (n=7) or (ii) multiplex ligation-dependent probe amplification (MLPA) (Probemix P335; MRC Holland, Amsterdam, the Netherlands) performed for this study to confirm findings or resolve discrepancies (n=29).

### Gene Expression by targeted RNA NGS

The normalized expression of a gene was calculated by summing unique RNA reads derived from all GSP2s targeting the gene and dividing by total number of unique RNA reads in the sample. Read counts were provided by AA output. No adjustments were made for the number of GSP2s, length of a gene, or batch effects. t-distributed stochastic neighbor embedding (t-SNE) plots were generated from log-transformed normalized expression using the R package Rtsne with perplexity set to 5 and default settings for other parameters. Annotations were based on available data from karyotype, FISH, FPH, RHP, OP, wtRNAseq, and low density arrays (LDA).

## Results

### Outlier analysis identifies known and novel *IKZF1*, *ERG*, and *PAX5* isoform junctions with high specificity for B-ALL

To investigate utility of aberrant isoforms, we screened for outlier expression of *IKZF1*, *ERG*, and *PAX5* isoforms in the heme cohort. By using an annotation-independent approach to maximize isoform recognition, we identified novel recurrent junctions (Fig 1) connecting *IKZF1* exons 2 or 3 to intergenic breakpoints (chr7:50498139, chr7:50505862, chr7:50525921) adjacent to putative cryptic splice acceptor sites approximately 25-50 kb downstream of *IKZF1*, thereby compatible with previously characterized DNA breakpoints associated with deletions involving exons 3-8 or 4-8^17,18^. Our approach similarly enabled detection of known pathogenic cryptic isoforms such as ERGalt despite their absence from annotation databases. Overall we identified outlier expression of 57 isoform junctions, with high specificity for a B-ALL component (96.5%; 193/200) and modest sensitivity for B-ALL (41.3%; 85/206) or MPAL with B-ALL (100%; 1/1) (SFig 1). A few non-specific (non B-ALL) cases narrowly exceeded outlier thresholds of common alternative splice junctions (*ERG* e6e8, *PAX5* e6e9), and were favored to reflect suboptimally low thresholds.

**Figure 1.**
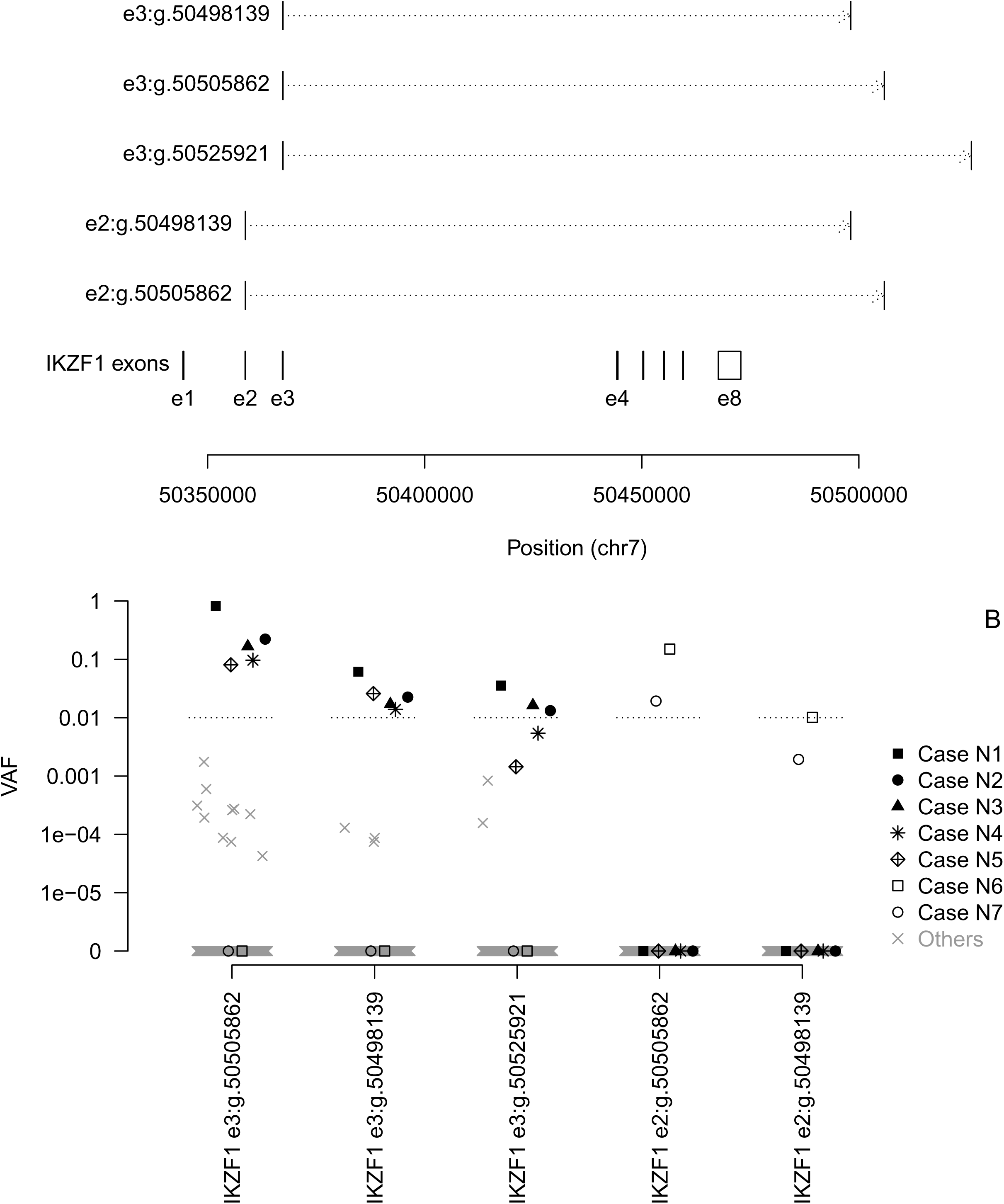
Novel recurrent *IKZF1* isoform junctions. (A) Our annotation-independent split-read approach identified novel junctions connecting *IKZF1* exons 2 or 3 to intergenic breakpoints approximately 25-50 kb downstream of *IKZF1*, thereby compatible with previously characterized DNA breakpoints associated with deletions involving exons 3-8 or 4-8^25,26^. Each of the intergenic breakpoints (chr7:50498139, chr7:50505862, chr7:50525921) was adjacent to AG putative cryptic splice acceptor sites. (B) B-ALL cases N1-N5 and N6-N7 expressed all three exon 3 and both exon 2 isoforms respectively and were associated with deletions of exons 4-8 and 3-8 respectively (Figure 2), thus compatible with alternative splicing where the chr7:50505862 splice site consistently had the dominant usage. All other cases had either no expression or very low level expression of at most one isoform, raising possibility of low-level read-through splicing.

### *IKZF1* exon-skipping isoforms at outlier expression levels are sensitive and specific markers for underlying intragenic and 3’ deletions

We compared RNA outlier expression status of *IKZF1* exon-skipping isoforms to DNA deletion status of the skipped exons (Figure 2, Table 1). We found outlier expression of the novel junctions e3:g.50505862 or e2:g.50505862 to be 100% sensitive and 100% specific for deletions of exons 4-8 (n=5) or exons 3-8 (n=2) respectively. Similarly, outlier expression of e1e4, e1e5 (without e1e4), e2e4, or e1e3 was 100% sensitive and 100% specific for uncommon deletions involving exons 2-3 (n=2), exons 2-4 (n=1), exon 3 (n=2), or exon 2 (n=1) respectively. Outlier expression of e1e8 was 100% sensitive (5/5) but 98.8% (256/259) specific for deletion of exons 2-7. Two of the false positives were associated with slightly different deletions (exons 1-7 and 2-8 instead of 2-7) incompatible as single events with formation of e1e8, thus raising the possibility of mischaracterized deletion boundaries or multiple deletions on separate alleles (e2-e7 on one allele combined with e1 or e8 on the other allele). Finally, outlier expression of e3e8, corresponding to the dominant negative isoform Ik-6, was the most prevalent aberration and was 92.9% (13/14) sensitive and 98.4% (246/250) specific for deletion of exons 4-7. The lone false negative had borderline RNA quality with an e3e8 VAF (0.93%) narrowly missing the outlier threshold (1.10%). The 4 false positives were favored to represent deletions of e4-e7 below the limit of detection of DNA-based assessment, given that RNA-based sensitivity for aberrant e3e8 was enhanced in 2 ways compared to DNA tests. First, background noise of e3e8 from presumed alternative splicing was minimal, resulting in a low outlier threshold (VAF 1.10%) well below the limit of detection by standard DNA testing for copy number events (typically VAF ≥ 10-20%). Second, RNA-based VAF of e3e8 was usually greater than DNA-based VAF of exon 4-7 deletions (p=0.00006 by paired t-test), empirically strengthening signal to noise and raising the possibility of allelic overexpression (Fig 3).

**Figure 2.**
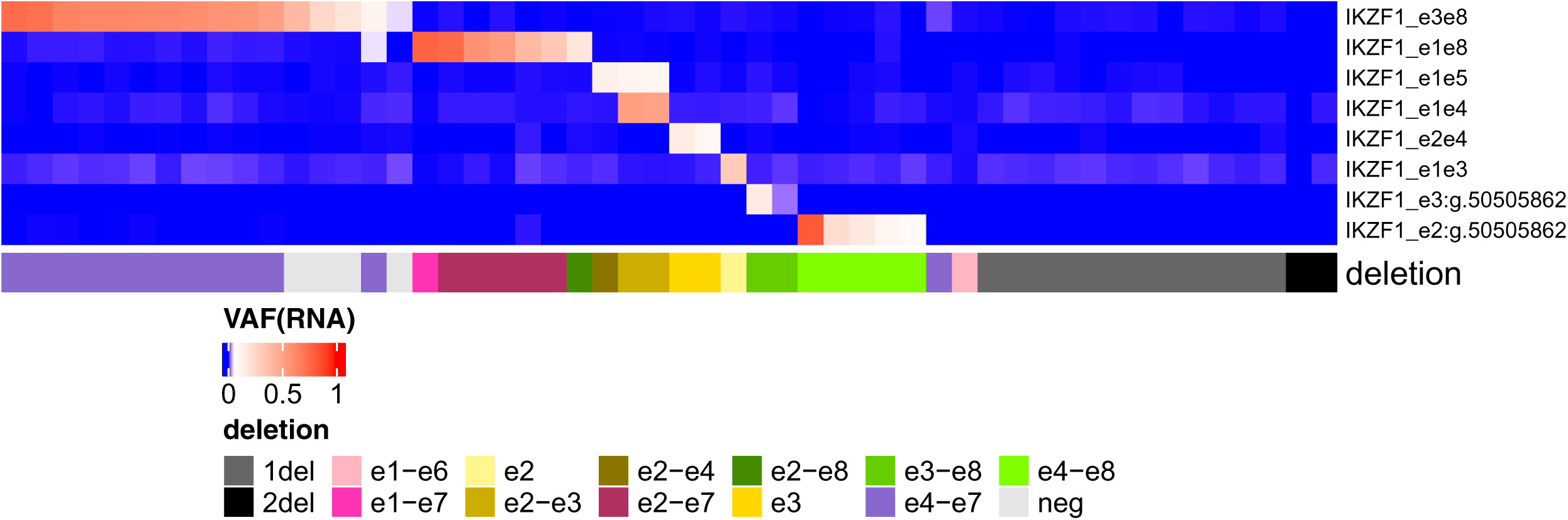
Outlier VAF of *IKZF1* isoforms versus *IKZF1* deletion status. Outlier VAF of eight *IKZF1* exon-skipping isoforms demonstrated an overall sensitivity of 97.0% (32/33) and specificity of 98.2% (226/231) for intragenic or 3’ deletion by DNA testing and almost always corresponded to deletion of the skipped exons. The false negative was associated with poor RNA quality; its VAF (0.93%) almost reached the outlier threshold (1.1%) and was higher than every other negative sample (next highest VAF of 0.44%), raising the possibility of a very low-level deletion (possibly from sampling differences) or a deletion with differential RNA degradation between tumor and wild-type cells. Of the 5 false positives, one was an e1e8 outlier associated with a slightly different but incompatible deletion (exons 1-7), thus favoring either incorrectly annotated deletion boundaries or multiple deletions on separate alleles (exon 2-7 and exon 1). The remaining 4 were e3e8 outliers favored to correspond to subclonal deletions below the limit of detection of DNA based methods (Figure 3).

**Figure 3.**
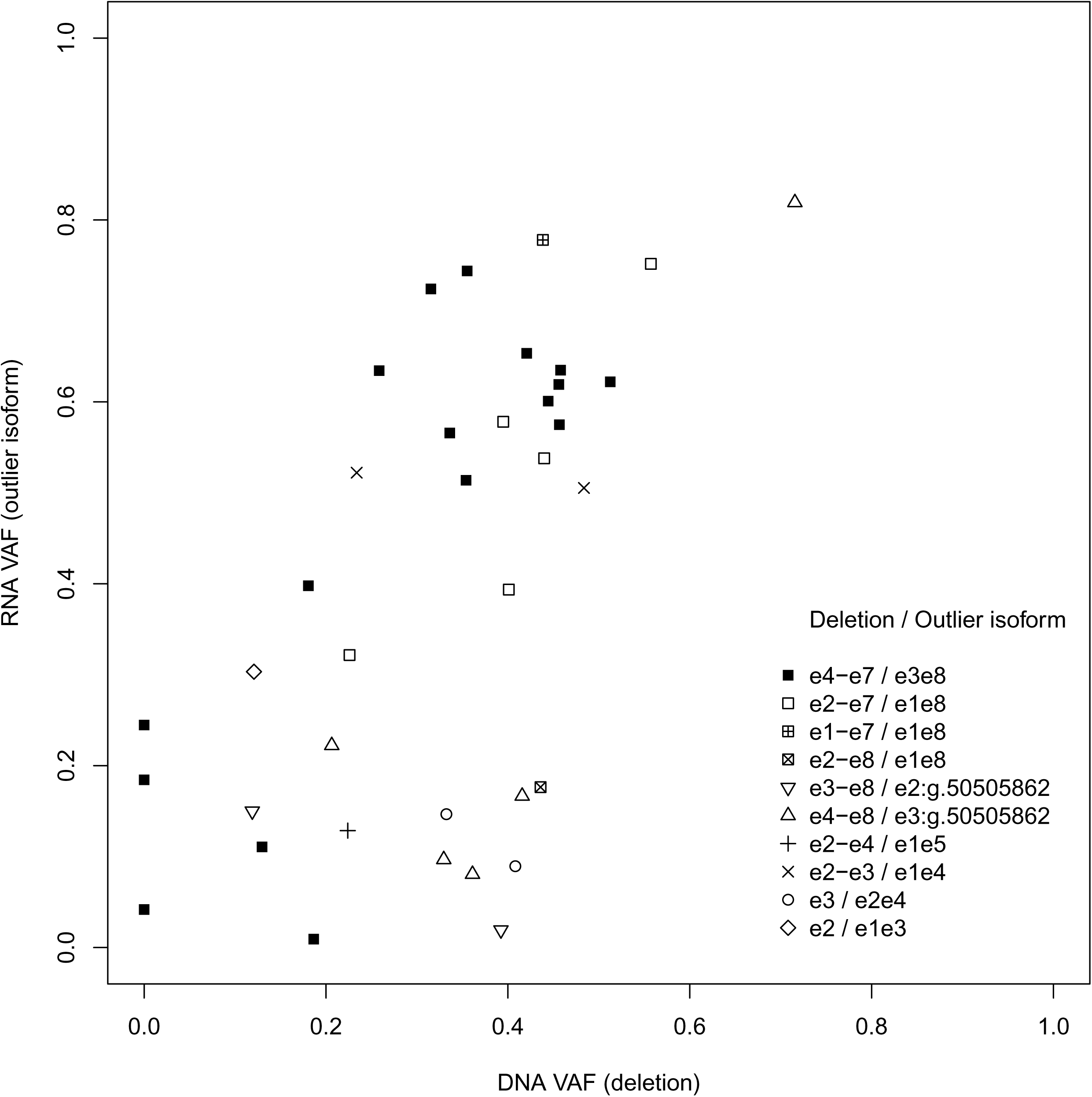
RNA-based VAF of *IKZF1* exon-skipping isoforms versus DNA-based VAF of deletions of the skipped exons. RNA-based VAFs of *IKZF1* e3e8 isoforms were consistently greater than DNA-based VAFs of the underlying deletions of exons 4-8, raising the possibility of RNA allelic overexpression of the mutant allele, however assay bias could not be entirely excluded. Regardless of the source, the behavior enhanced limit of detection by FPH empirically.

**Table 1.**
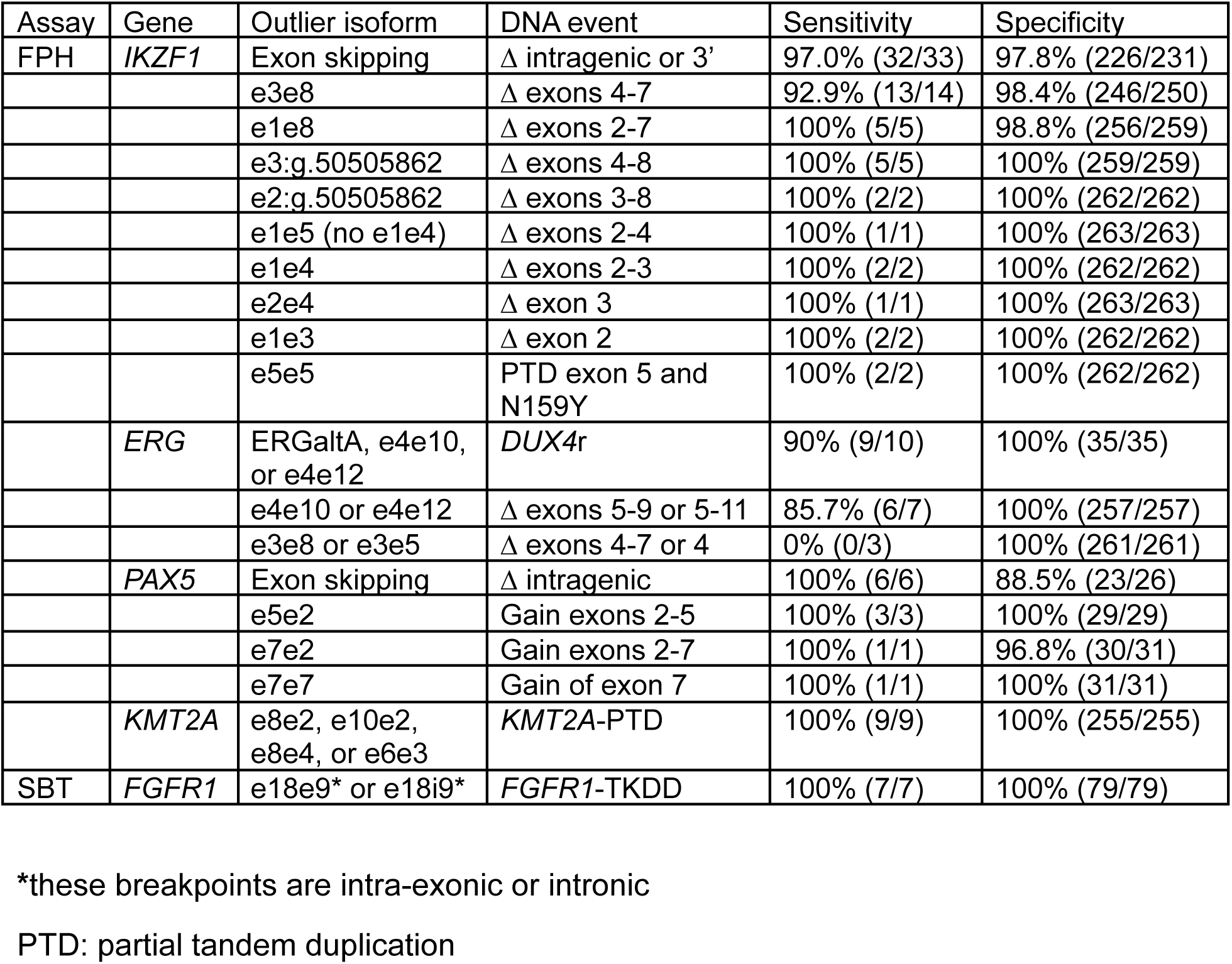
Outlier expression of isoforms as markers of DNA event.

We also explored whether whole gene or 5’ deletions of *IKZF1* might be detectable through haploinsufficient *IKZF1* gene expression but found no significant differences between B-ALL cases with shallow *IKZF1* deletions (p=0.36) versus non-deleted B-ALL (defined as having neither deletions nor outlier isoform status), although a trend appeared for deep *IKZF1* deletions (p=0.06) (SFig 2). Rather, we only found differential *IKZF1* gene expression in e3e8 outliers (p=0.0007; up-regulation) versus non-deleted B-ALL, again suggestive of allelic overexpression.

### B-ALL with *IKZF1* N159Y have recurrent *IKZF1*-PTD of exon 5 duplicating N159Y

We compared outlier status to molecular characteristics (SFig 1). In 2 of 2 B-ALL cases harboring the *IKZF1* N159Y mutation (a proposed emerging subtype with a distinct gene expression profile^5^), we identified outlier expression of *IKZF1* e5e5 (VAFs 0.443-0.517) compared to zero expression in virtually all remaining cases (396/397), except for 1 case with 1 e5e5 read (VAF 0.006). *IKZF1* exon 5 was not targeted by FPH, arguing against circRNA of *IKZF1* exon 5 as a source of e5e5 reads. Rather, we found e5e5 reads in both cases derived from all 5 *IKZF1* primers (targeting forward strands of exons 1-3 and reverse strands of exons 7-8), consistent with mRNA transcripts extending from exon 1 to exon 8 with PTD of exon 5. Since *IKZF1* N159Y is located within exon 5, we asked whether N159Y or PTD occurred first or independently in clonal evolution. By identifying e5e5 reads with N159Y on either side of the e5e5 junction, we deduced PTD as the secondary event in both cases (SFig 3). RHP did not report copy number gains of exon 5, thus we manually reviewed copy number data and raw RHP alignments over *IKZF1*, revealing small partial tandem duplications (PTD) of sizes 344 bp and 396 bp spanning *IKZF1* exon 5 and duplicating N159Y (Fig 4). Given this unexpected finding, we screened the entire historical RHP cohort. We did not find PTD of exon 5 in multiple cases with N159S/T, and there were no further instances of N159Y. However, among AML cases without N159 variants, we identified very rare in-frame ITDs within exon 5 spanning the N159 codon.

**Figure 4.**
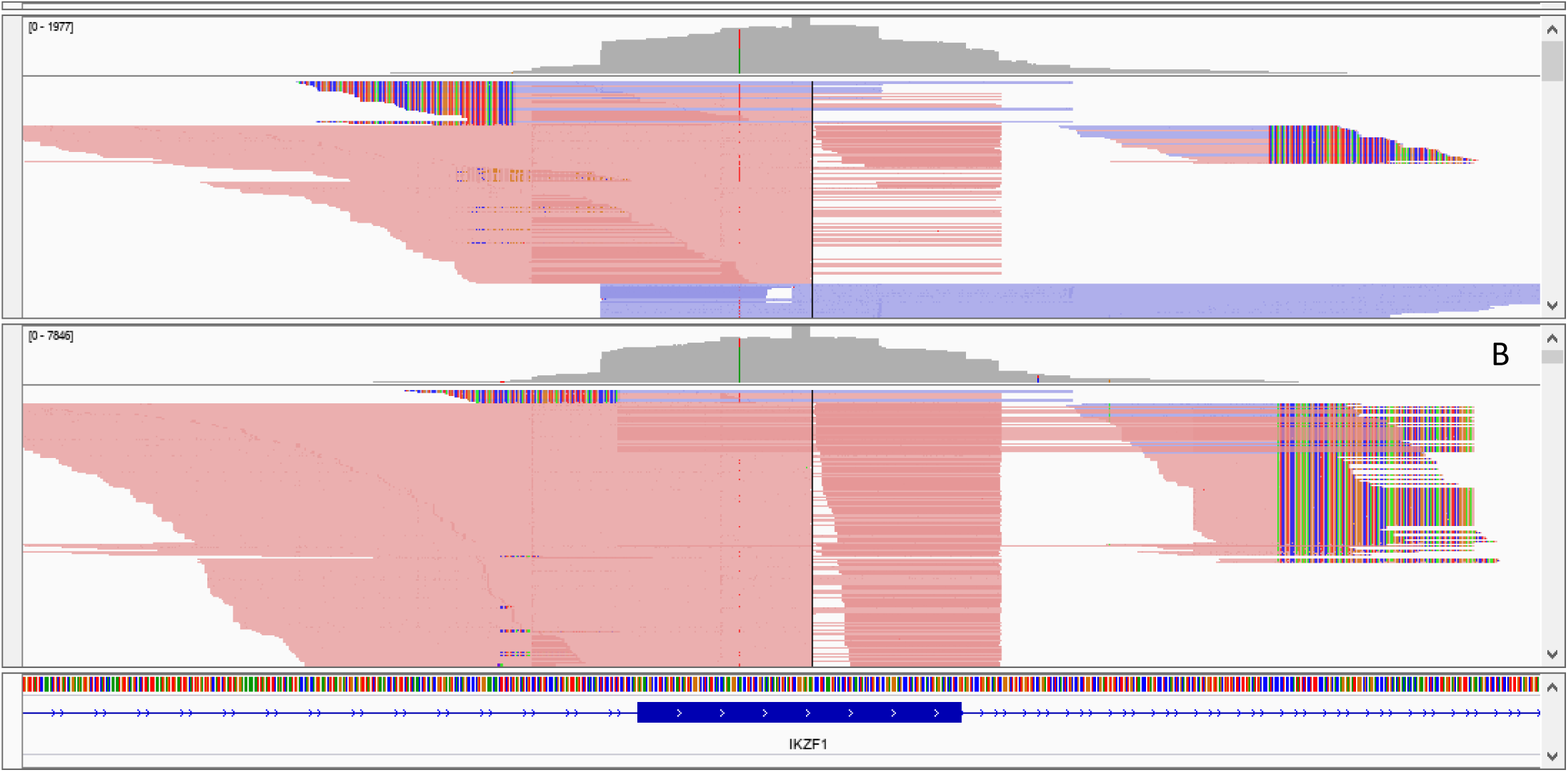
*IKZF1*-PTD of exon 5 in B-ALL with N159Y. In 2 of 2 B-ALL cases harboring *IKZF1* N159Y variants, manual review in IGV revealed partial tandem duplications spanning exon 5 with breakpoints in introns 4 and 5 and with lengths (A) 344 bp and (B) 396 bp, corresponding to RNA outlier expression of *IKZF1* e5e5. The breakpoints were sufficiently near exon 5 to be captured fortuitously by RHP baits targeting exon 5 but were undetected by the informatic pipeline due to lack of a structural variant caller and filtering of intronic variants regardless. In both cases, N159Y was present 5’ of the mutant junction in some reads and 3’ of the mutant junction in other reads, implying duplication of N159Y occurring after N159Y in the clonal hierarchy. The PTDs were described by the following modified notation: A. c.589+165_589+166insA/422-10_589+165[D1:c.475A>T;D2:c2.475A>T] B. c.589+160_589+161insTCAC/422-64_589+160[D1:c.475A>T;D2:c2.475A>T] We also manually reviewed the copy number profiles from RHP, which by convention did not report single exon changes due to noise, however we did not observe gain of exon 5 in either case. The small PTDs may have contributed to the absence of a copy number signal due to loss of supplementary alignment read counts in the CNV algorithm and disrupted UMI reduction in the UMI algorithm.

In contrast to *IKZF1* e5e5 and its association with PTD of exon 5, our screening approach yielded 2 samples (1 B-ALL and 1 eosinophilia) with outlier expression of *IKZF1* e3e2 minimally exceeding thresholds, where the pattern of originating primers of e3e2 reads was consistent with circRNA of *IKZF1* e2-e3 and not PTD of exons 2-3. Namely, e3e2 reads were derived almost exclusively from primers targeting forward strands of exons 2-3 whereas significant fractions of e3e4 reads were derived from primers targeting each of the exons 1-3.

### Outlier expression of either ERGalt A or *ERG* isoforms is highly sensitive and specific for *DUX4*-rearranged B-ALL

We investigated the utility of outlier *ERG* isoforms as markers for *DUX4*r B-ALL. We found that outlier expression of either (i) ERGaltA (threshold of 11.0% VAF) or (ii) *ERG* e4e10 or e4e12 (thresholds of 1%) was 90% (9/10) sensitive and 100% (35/35) specific for *DUX4*r status determined by wtRNAseq (Fig 5). Both criteria (i) and (ii) contributed to optimizing performance since their sensitivities as standalone markers were 80% and 50% respectively. We identified outlier expression in 8 additional cases without wtRNAseq data; all were B-ALLs lacking an established molecular subtype, including 2 with intragenic *ERG* deletions by DNA NGS and 1 with a DUX4/ERG signature by LDA screening. Among the overall cohort, cases with deletions of *ERG* exons 5-9 or 5-11 were nearly always (86%; 6/7) associated with outlier e4e10 or e4e12 expression, where the case lacking isoform expression was complicated by borderline RNA quality (same false-negative case with low-level outlier *IKZF1* e3e8); by contrast, cases with deletions of *ERG* exons 4-7 (n=2) or exon 4 (n=1) had zero expression of corresponding isoforms e3e8 or e3e5.

**Figure 5.**
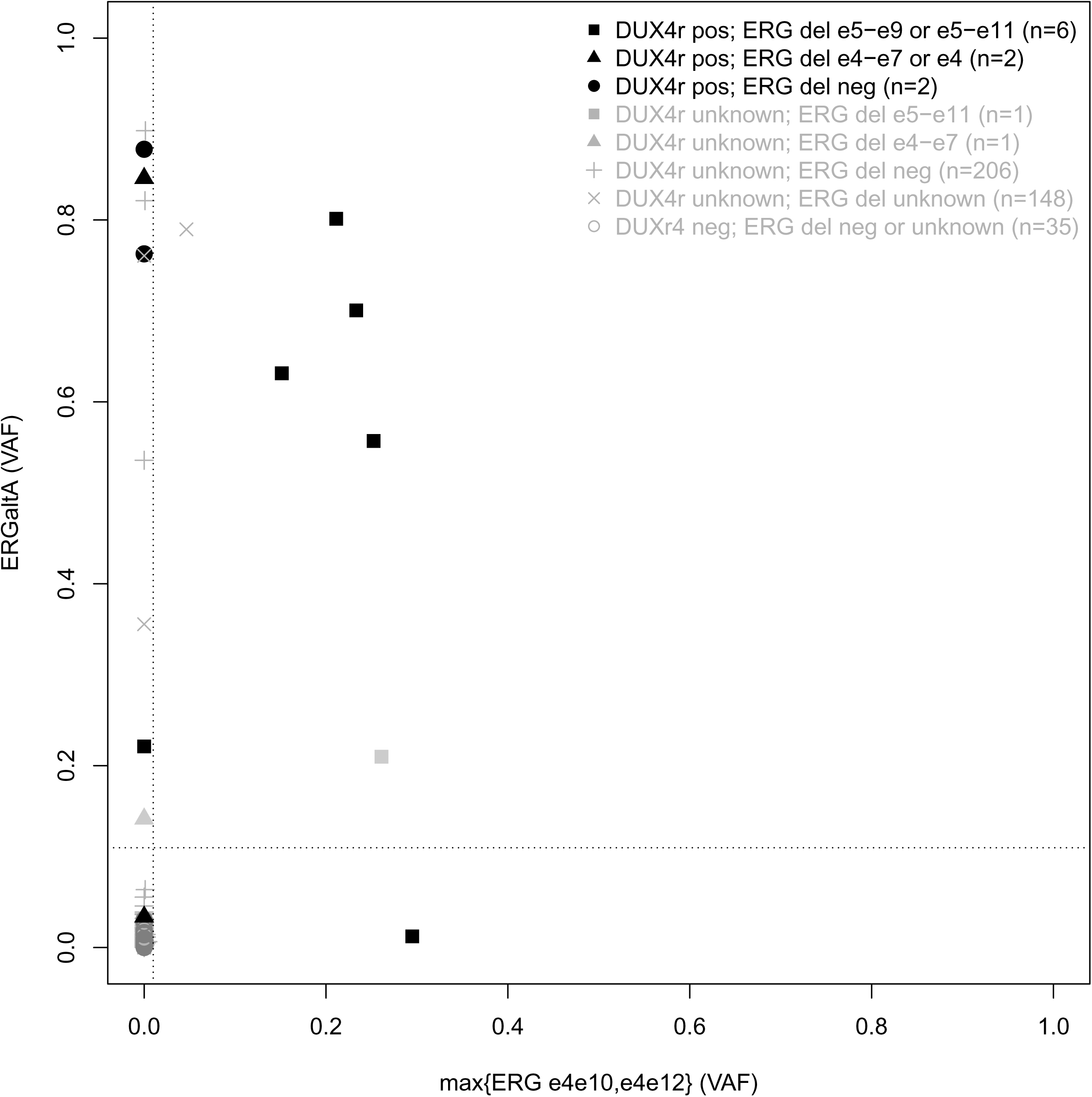
Outlier VAF of ERGaltA, *ERG* e4e12, or *ERG* e4e10 versus *DUX4* rearrangement status and *ERG* deletion status. Outlier expression of either ERGaltA, *ERG* e4e10, or *ERG* e4e12 was 90% (9/10) sensitive and 100% (35/35) specific for *DUX4* rearrangements as determined by wtRNAseq. Outlier expression of e4e10 or e4e12 was 85.7% (6/7) sensitive and 100% (249/249) specific for intragenic deletions of exons 5-9 or 5-11, where the lone false negative had borderline RNA quality. By contrast, there was zero expression of e3e8 or e3e5 in cases with deletions of exons 4-7 or exon 4.

We observed high *PDGFRA* gene expression in the lone false negative case not identified by outlier *ERG* analysis. *PDGFRA* overexpression has been associated with *ERG* deletions and binding of DUX4 to the *PDGFRA* transcription start site in *DUX4*r^1,19^, however we also observed high expression in a *DUX4*r-negative B-ALL in our cohort, limiting its specificity. Given the known distinct gene expression profile of *DUX*4r B-ALL, we analyzed for differential gene expression among the 100 genes targeted by FPH, yielding 8 genes with significant overexpression in *DUX4*r-positive versus *DUX4*r-negative B-ALL. These genes demonstrated at most 30% sensitivity to achieve 100% specificity for *DUX4*r, thus ERGaltA had the top performance as a single positive marker for *DUX4*r (80% sensitivity, 100% specificity). Similarly, tSNE relative to FPH gene expression data generated a cluster consisting predominantly but not exclusively of *DUX4*r (SFig 4).

### PAX5amp is associated with *PAX5* out-of-order isoforms at high VAFs

We screened *PAX5* isoforms harboring out-of-order exons at outlier levels for potential PAX5amp candidates. We observed robust background expression of e5e2 across B-ALL cases, compatible with known circRNA involving *PAX5* exons 2-5 and resulting in a relatively high outlier threshold. We identified 4 cases (P1-P4) with clear outlier expression of e5e2 (67.7% VAF), e5e2 (21.8%), joint e7e2 (55.0%) and e5e2 (20.5%), and e7e7 (21.5%) respectively (Fig 6A). Using MLPA, we confirmed parsimonious underlying *PAX5* copy number alterations characterized by (P1) high level gain of exons 2-5, (P2) shallow gain of exons 1-5 (compatible with e5e2 isoforms since e1 lacks a splice acceptor to enable e5e1 splicing), (P3) several different copy number levels suggesting multiple gain events involving exons 1-7, 2-7, and 2-5, and (P4) shallow gain of exon 7 (Fig 6B-E). The cases P1 and P3, with high RNA VAFs above 50% and high level copy number gains, lacked an alternative molecular subtype, arguing for the emerging PAX5amp subtype, whereas P2 harbored an ABL class fusion (*PDGFRB*), which has been previously described in this context^5^, and P4 had a clonal *PCF3-TBX1* rearrangement (SFig 5). Two additional cases narrowly exceeded outlier thresholds of e7e2 and e4e2 and were favored to represent circRNAs and suboptimal thresholds; the e4e2 case was associated with a *PAX5* fusion with an exon 4 breakpoint, suggesting altered circRNA formation in the context of rearrangement (Fig 6A, SFig 5). Limited MLPA and microarray testing of cases without outlier expression of out-of-order *PAX5* isoform junctions identified no other instances of intragenic *PAX5* gains (n=28).

**Figure 6.**
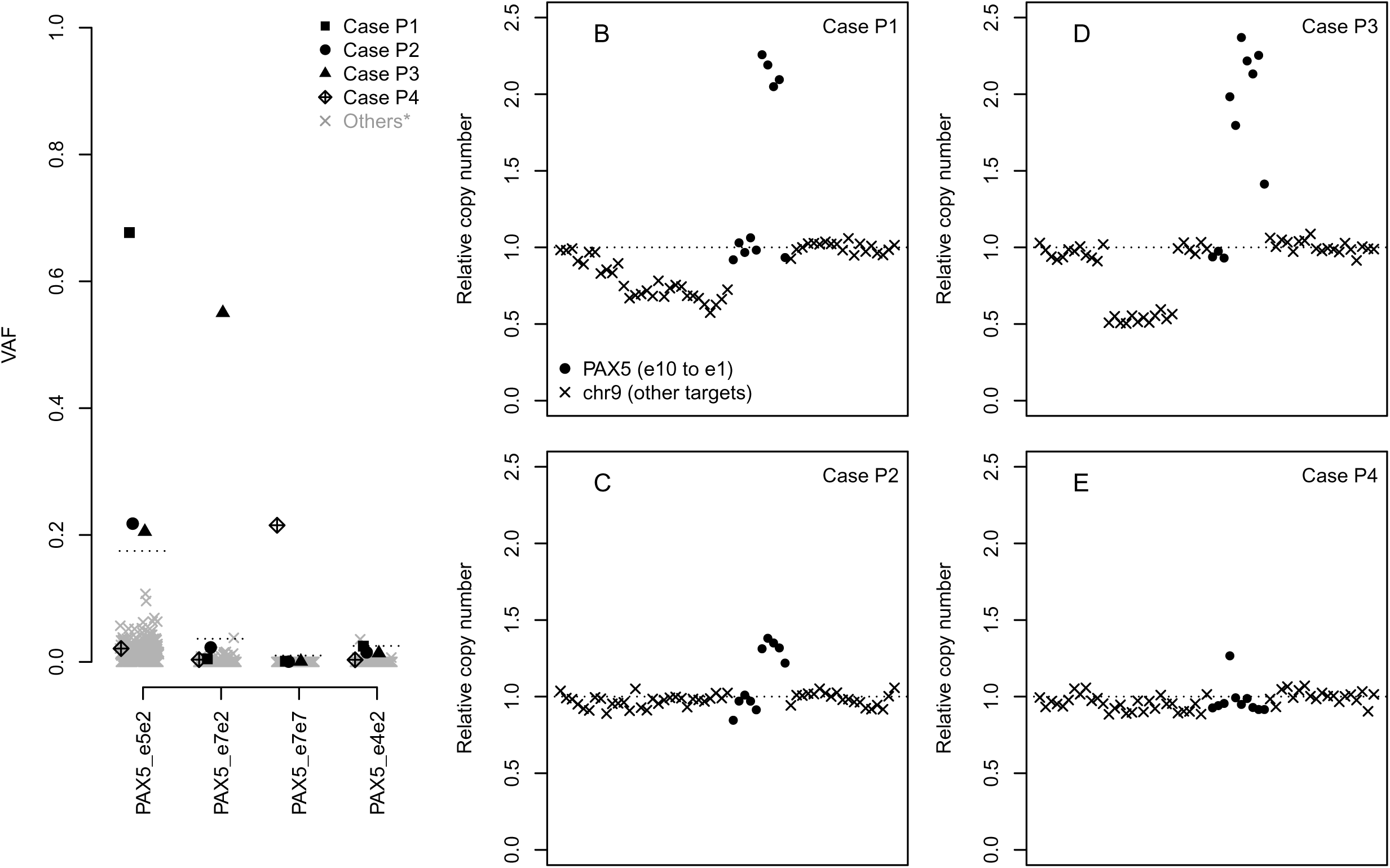
Outlier expression of *PAX5* out-of-order exon junctions may be sensitive but not specific for PAX5amp. (A) Isoform expression. Four cases (P1-P4) had clear outlier expression of *PAX5* e5e2, e7e2, and/or e7e. Another case minimally exceeded the outlier threshold for *PAX5* e4e2 and harbored a *PAX5* rearrangement involving exon 4, thus favoring altered circRNA expression in the context of a fusion and a suboptimally low threshold. A final case, which barely exceeded the outlier threshold for *PAX5* e7e2, was similarly favored to reflect circRNA and a suboptimal threshold. (B-E) Copy number gains by MLPA. Cases P1 (B) and P3 (D) demonstrated higher order intragenic gains within *PAX5* by MLPA, were not associated with a known molecular subtype, and were favored to represent PAX5amp. P3 demonstrated 4 probably copy number number levels in *PAX5* suggestive of multiple gain events (e1-e7, e2-e7, and e2-e5), thereby consistent with the high outlier e7e2 and moderate outlier e5e2 VAFs. Cases P2 (C) and P4 (E) demonstrated shallow intragenic gains and harbored other rearrangements determining their molecular subtypes

### *PAX5* exon-skipping isoforms at outlier levels are associated with intragenic deletions and facilitate specific but not sensitive detection of the *IKZF1*^plus^ signature

We next screened exon-skipping *PAX5* isoforms for potential intragenic deletions (SFig 5–6). We identified outlier expression across 30 samples involving 11 junctions (e1e6, e1e7, e1e8, e1e9, e5e7, e5e8, e5e9, e5e10, e6e8, e7e9, e8e10). Outlier e5e8, e5e9, e7e9, or e8e10 frequently co-occurred with *CRLF2* rearrangements, while outlier e1e7, e1e8, or e1e9 frequently co-occurred with *ETV6-RUNX1*, as has been previously described (SFig 5)^20^. We observed single exon-skipping isoforms (e7e9, e8e10) at high outlier VAFs (>85%) favoring loss of heterozygosity in 3 cases, and outlier co-expression of multiple isoforms similarly raising the possibility of biallelic dysfunction in 2 other cases (Q4-Q5). We compared outlier expression to intragenic deletions in a limited cohort (n=32) and found outlier expression of e1e7, e7e9, e5e8, e5e9, or e5e10 to be 100% (6/6) sensitive (cases Q1-Q6) and 88.5% (23/26) specific for intragenic deletions. The 3 false positives (Q1-Q3) all had low outlier VAFs (< 20%) and were suspected to represent deletions below the limit of detection of DNA testing, similar to *IKZF1* findings. The 2 cases (Q4-Q5) with outlier co-expression of multiple isoforms demonstrated complicated copy number profiles implying multiple intragenic deletion events.

We evaluated for the B-ALL high-risk *IKZF1*^plus^ signature, defined by *IKZF1* deletion combined with one or more deletions of *PAX5*, *CKDN2A*, *CDKN2B*, and/or the PAR1 region in the absence of *ERG* deletion^7^. We used outlier expression of *IKZF1*, *PAX5*, and *ERG* exon-skipping isoforms as makers of focal deletions, and used the fusion *P2RY8*-*CRLF2*, whose detection was clinically validated under FPH, as a proxy for PAR1 deletion. FPH accordingly identified 10 B-ALL cases with focal *IKZF1* deletions satisfying *IKZF1*^plus^, composed of 4 with inferred *PAX5* intragenic deletions alone, 4 with *P2RY8-CRLF2* alone, and 2 with both. Sensitivity for *IKZF1*^plus^ was limited by inability to detect broad *IKZF1*, *PAX5*, and *CDKN2A* deletions and a lack of *CDKN2B* targets.

### Outlier expression of *KMT2A* e8e2, e10e2, e8e4, or e6e3 is sensitive and specific for the corresponding *KMT2A*-PTD at initial diagnosis

We tested the ability of outlier isoform expression to detect *KMT2A*-PTD as determined by clinically validated orthogonal testing, where the known *KMT2A*-PTD cases (1 pediatric, 7 adult, and the cell line EOL1) corresponded to the isoforms e8e2 (5), e10e2 (2), e8e4 (1), and e6e3 (1). Positive (n=9) and negative (n=255) *KMT2A*-PTD cases were completely separated by outlier expression thresholds (Figure 7A). Rare non-PTD cases narrowly exceeded VAF cutoffs for e8e2 but remained below the normalized expression cutoff (Figure 7B); these occurred in *KMT2A*-rearranged leukemias involving exon 8, consistent with altered circRNA formation as described previously^21^. The *KMT2A* junction e8e4 had the highest background expression, presumably derived from the known circRNA species hsa_circ_0095112^22^. By contrast, *KMT2A* junction e6e3 was expressed in only 2 non-PTD cases at 1 split-read.

**Figure 7.**
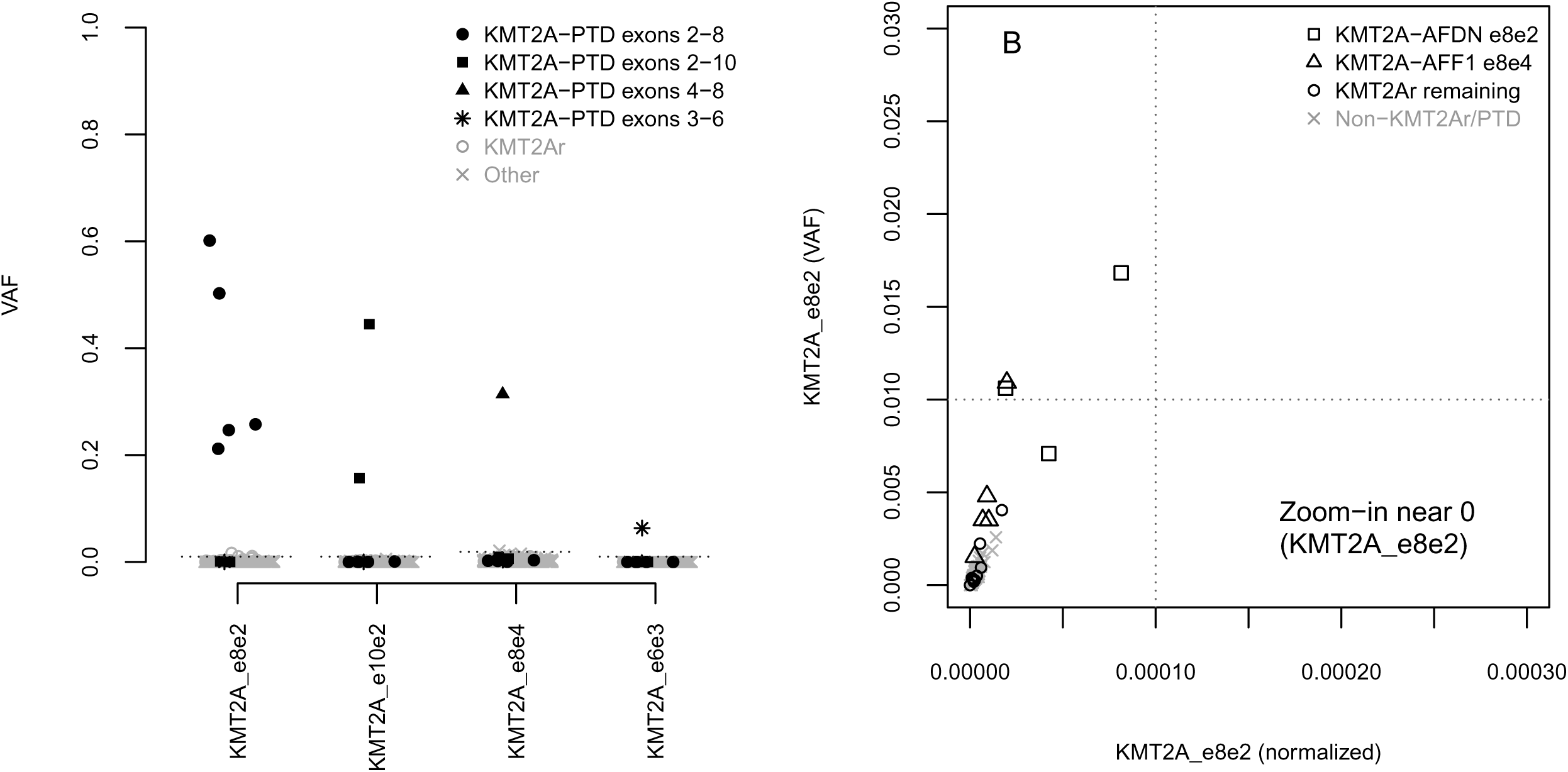
*KMT2A* out-of-order exon junctions as a marker of *KMT2A*-PTD. Outlier expression of *KMT2A* e8e2, e10e2, e8e4, or e6e3 was 100% (9/9) sensitive and 100% (255/255) specific for known *KMT2A*-PTDs of exons 2-8, 2-10, 4-8, or 3-6. The highest VAFs suggested the presence of allelic complexity, such as copy neutral loss of heterozygosity, higher order duplications, or episomal amplifications. Note also that our definition of expressed VAF underestimated the true percentage of *KMT2A*-PTD transcripts since PTD transcripts also contained wild-type junctions causing an overestimated wild-type denominator; the true fraction of *KMT2A*-PTD transcripts was more accurately assessed by the formula 1/((1/VAF)-1). The highest expression levels of e8e2 after *KMT2A*-PTDs occurred in *KMT2A*r cases with fusion breakpoints involving *KMT2A* exon 8, however these never reached outlier status. *KMT2A* wild-type cases occasionally demonstrated a few e8e2 reads, consistent with infrequent low-level circRNA.

### Split-read analysis of *FGFR1* breakpoints spanning the kinase domain is sensitive and specific for *FGFR1*-TKDD

We applied outlier analysis to *FGFR1*-TKDD. In brain tumor samples tested by SBT, the detection of split-reads with a 5’ breakpoint within exon 18 and a 3’ breakpoint within exon 9 or intron 9 was 100% (7/7) sensitive and 100% (79/79) specific for detection of *FGFR1-TKDD* relative to Oncopanel, where the positive cases were all gliomas. RNA mutant breakpoints under SBT agreed with genomic DNA breakpoints under Oncopanel in all positive samples. No solid tumor samples (non-brain) demonstrated split-reads characteristic of *FGFR1*-TKDD. Given the fact that breakpoints in exon 18 were intraexonic and not adjacent to potential cryptic splice sites, low level expression from alternative sources such as circRNA was never present.

## Discussion

Contemporary B-ALL risk stratification utilizes *IKZF1* deletion status to improve treatment outcomes^23^. In FPH, we found outlier expression of known and novel *IKZF1* exon-skipping isoforms to be effective markers of intragenic and 3’ deletions with 97.0% (32/33) sensitivity and 97.8% (226/231) specificity overall. Moreover, the false negative was associated with suboptimal RNA quality, and the false positives were either suspected to represent subclonal deletions below the limit of detection of our DNA-based assays or were due to minor discrepancies in deletion boundaries. Targeted RNA sequencing may accordingly be more reliable than wtRNAseq, which has successfully detected deletions of exons 4-7 through the e3e8 isoform but has been less successful with other deletions including exons 4-8^13,24^. In FPH, accurate quantification and optimal distinction from background noise may have benefited from direct targeting of relevant junctions, deeper coverage, and use of unique molecular identifiers. Alternatively, it is possible that wtRNAseq performance may improve from different informatic approaches, including the outlier methods used here or the recently developed MINTIE algorithm^25^. However, the MINTIE study reported a limited number of cases (n=7) with *IKZF1* RNA variants and may not have had orthogonal DNA data to assess performance. Thus, head-to-head comparisons with uniform informatics of targeted RNA sequencing and wtRNAseq may be informative.

In FPH, RNA VAFs of e3e8 were significantly greater than DNA VAFs of exon 4-7 deletions, suggesting allelic overexpression and empirically improving limit of detection regardless of whether mediated biologically or by assay bias; this behavior might also partially explain the successful use of e3e8 in wtRNAseq studies. More generally, aberrant isoforms across genes frequently demonstrated minimal background noise and defaulted to 1% outlier thresholds, potentially enabling subclonal detection of underlying DNA events well below the limit of detection of standard DNA-based assays^26,27^. Copy number changes involving a single exon present another challenging category for DNA-based assays, since their signal may be identical to noise, whereas outlier isoform analysis can more reliably ensure the high specificity required for clinical diagnostics. We identified unrecognized recurrent PTDs of *IKZF1* exon 5 duplicating N159Y on both the RNA and DNA levels in the B-ALL subtype with mutated N159Y. RNA e5e5 reads were similarly detected by MINTIE in a B-ALL with the gene expression signature of *IKZF1* N159Y, although mutational status was not described^25^. The significance of PTD and its contribution to the distinct gene expression profile associated with N159Y will require further study.

Robust isoform detection based on systematic outlier analysis importantly facilitates recognition of new genetically defined subtypes of B-ALL specified by the International Consensus Classification and the 5th edition of the WHO, thereby complementing fusion detection to enable more complete molecular classification within a single targeted RNA sequencing assay^28^. We found outlier expression of *ERG* isoforms to be 90% (9/10) sensitive and 100% (35/35) specific for the relatively frequent *DUX4*r subtype, which has been underrecognized historically since the rearrangements involve cytogenetically cryptic small insertions that are also difficult to molecularly target due to the location of *DUX4* within a D4Z4 macrosatellite repeat array^30^. Performance of these *ERG* isoform markers has been less strong in wtRNAseq^1,2^, similar to *IKZF1*. False negatives in FPH may be rescued by gene expression screening, however *DUX4*r candidates identified in this way should be confirmed by orthogonal methods due to imperfect specificity of gene expression analyses. FPH may also be capable of detecting the complete spectrum of *PAX5* variants (P80R, rearrangements, and intragenic amplifications) defining the B-ALL with mutated *PAX5* P80R subtype and the provisional B-ALL with *PAX5* alteration subtype. PAX5amp e5e2 transcripts must be distinguished from circRNA of *PAX5* exons 2-5, which is upregulated in B-cells and captured by assays lacking a polyA mRNA enrichment step ^31,32^. In our cohort, e5e2 junctions were present in 98.7% of B-ALL samples, highlighting the importance of accurate quantification.

Outlier analysis is most effective for genomic events that yield robust expression of RNA isoforms but may be challenging for poorly expressed genes. Our systematic rule for defining outlier thresholds was also relatively arbitrary, and performance may be improved by selective isoform-specific modifications. Overall however, we find that properly defined outlier analysis of RNA isoforms can be used to identify multiple clinically relevant genomic variants, and hypothesize that this analysis can be extended to additional genomic loci of clinical significance.

## Data Availability

Data produced in the present study are available upon reasonable request to the authors.

## Acknowledgements

This work was supported by a St. Baldrick’s Foundation Consortium grant, Charles H. Hood Foundation Grant (YP), NIH K08 CA222684 (YP), the clinical research staff of Dana-Farber Cancer Institute (Asmani Adhav, Hannah McLane), the study team of the DFCI ALL Consortium Protocol 16-001, the staff of the Center for Advanced Molecular Diagnostics at Brigham and Women’s Hospital, and the staff of the Laboratory for Molecular Pediatric Pathology at Boston Children’s Hospital. We also thank the lab of Benjamin Ebert for the gift of EOL-1 cells.

## Author Contributions

HKT and MHH designed the study and wrote the manuscript. THT, YP, ASK, VN, LBS, and MHH contributed data. VL, JT, YL, and LG performed experimental work. HKT developed informatic tools. All authors performed data analysis and edited the manuscript.

## Supplementary Figure Legends

**SFig 1.**
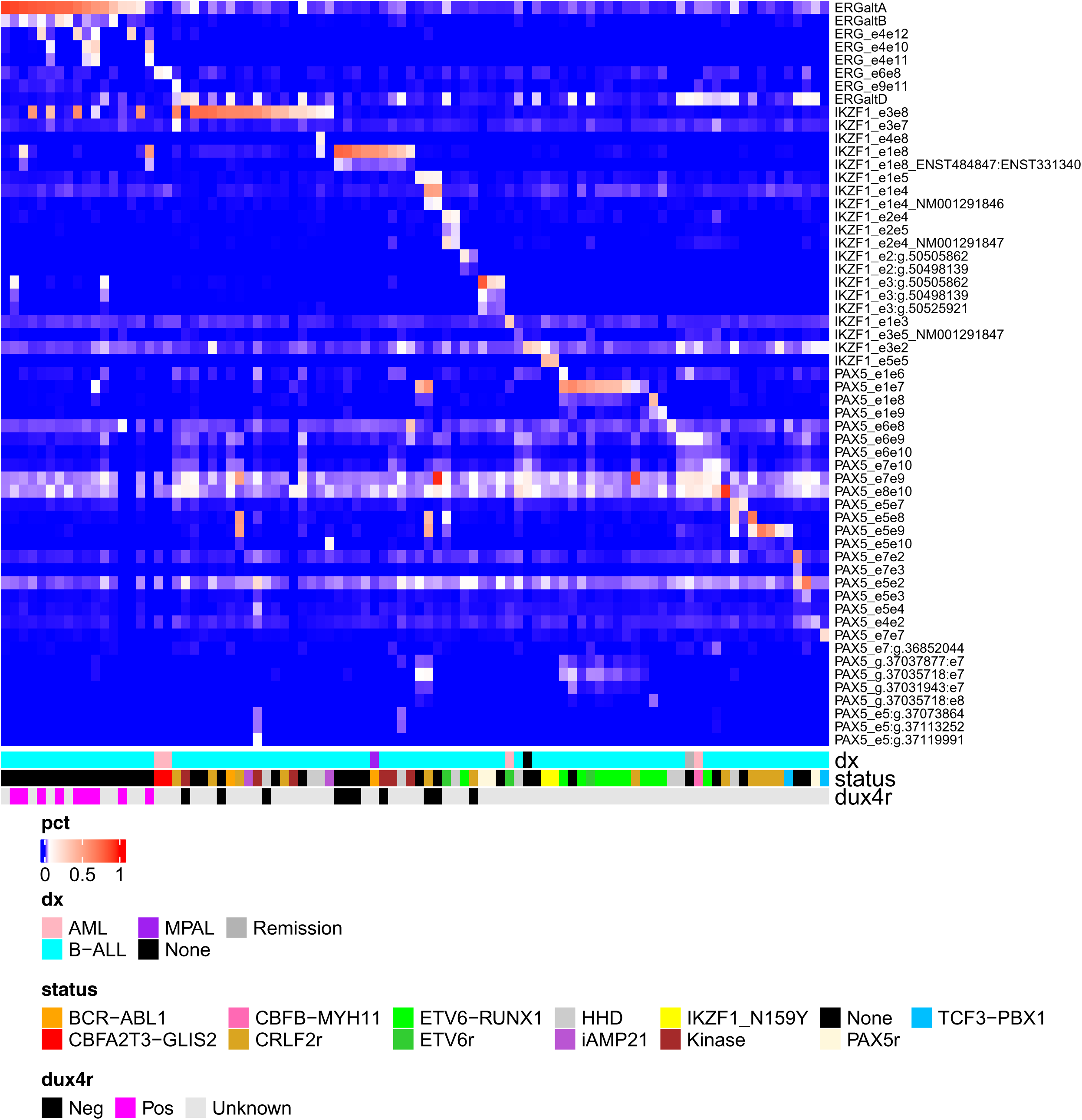
Outlier expression of *IKZF1*, *ERG*, or *PAX5* isoforms. In the heme cohort (n=407), outlier expression occurred for 57 isoform junctions across 92 samples consisting of 85 B-ALLs, 1 *BCR-ABL1*-positive MPAL (characterized by a predominant population of B-ALL blasts and a minor population of AML blasts), 4 AMLs (including 2 serial samples from the same patient), 1 remission sample, and 1 sample without a neoplastic diagnosis, thus demonstrating an overall 41.5% (86/207) sensitivity and 96.5% (193/200) specificity for a component of B-ALL. The serial AMLs had outlier expression of *ERG* e6e8 barely exceeding the outlier threshold, suggestive of false positives from alternative splicing and a suboptimally low threshold. Other isoform junctions with lower expression levels were highly correlated with expression of another isoform, consistent with alternative splicing in the context of the same underlying DNA structural event; these included *IKZF1* e1e4 (NM001291846), e1e8 (ENST0000484847:ENST0000331340), e2e5, e2e4 (NM001291847), e3:g.50498139, e3:g.50525921, and e2:g.50498139, ERGaltB and *ERG* e4e11, and *PAX5* e5e3 and e5e4. Comparisons with established and emerging B-ALL molecular subtypes showed various associations including (i) *IKZF1* e5e5 with *IKZF1* N159Y (Fig 4, SFig 3), ERGaltA, *ERG* e4e10, or *ERG* e4e12 with *DUX4*r (Fig 5), and certain *PAX5* deletions with *CRLF2*r or *ETV6-RUNX1* (SFig 5).

**SFig 2.**
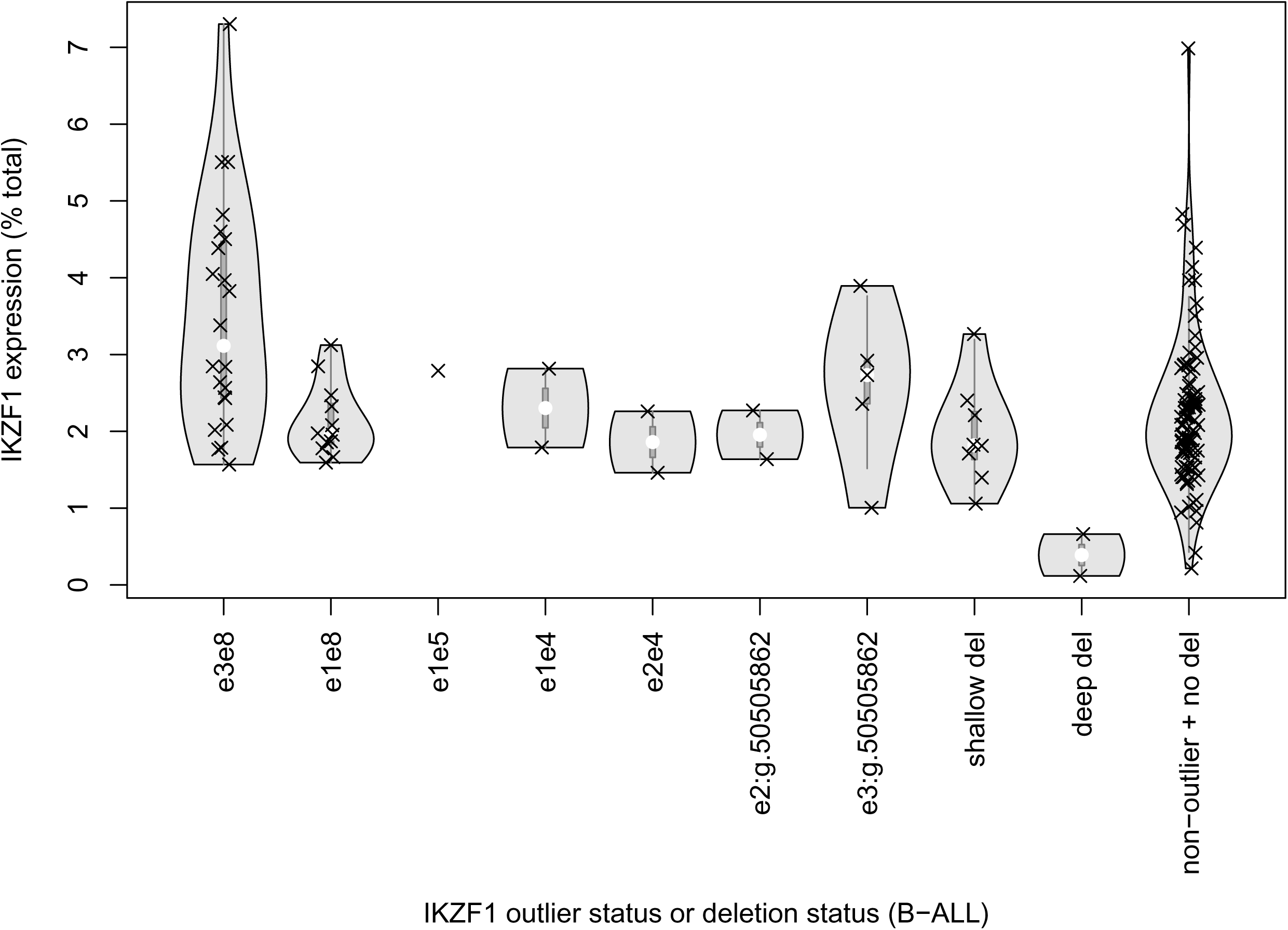
*IKZF1* gene expression does not reliably distinguish haploinsufficiency. Cases with broad *IKZF1* shallow (single-copy) deletions were not distinguished from non-deleted B-ALL based on *IKZF1* gene expression under FPH (p=0.36), although a trend appeared for deep *IKZF1* deletions (p=0.06). *IKZF1* gene expression was significantly higher in cases with e3e8 outliers versus non-deleted B-ALL (p=0.0007), raising the possibility of allelic overexpression.

**SFig 3.**
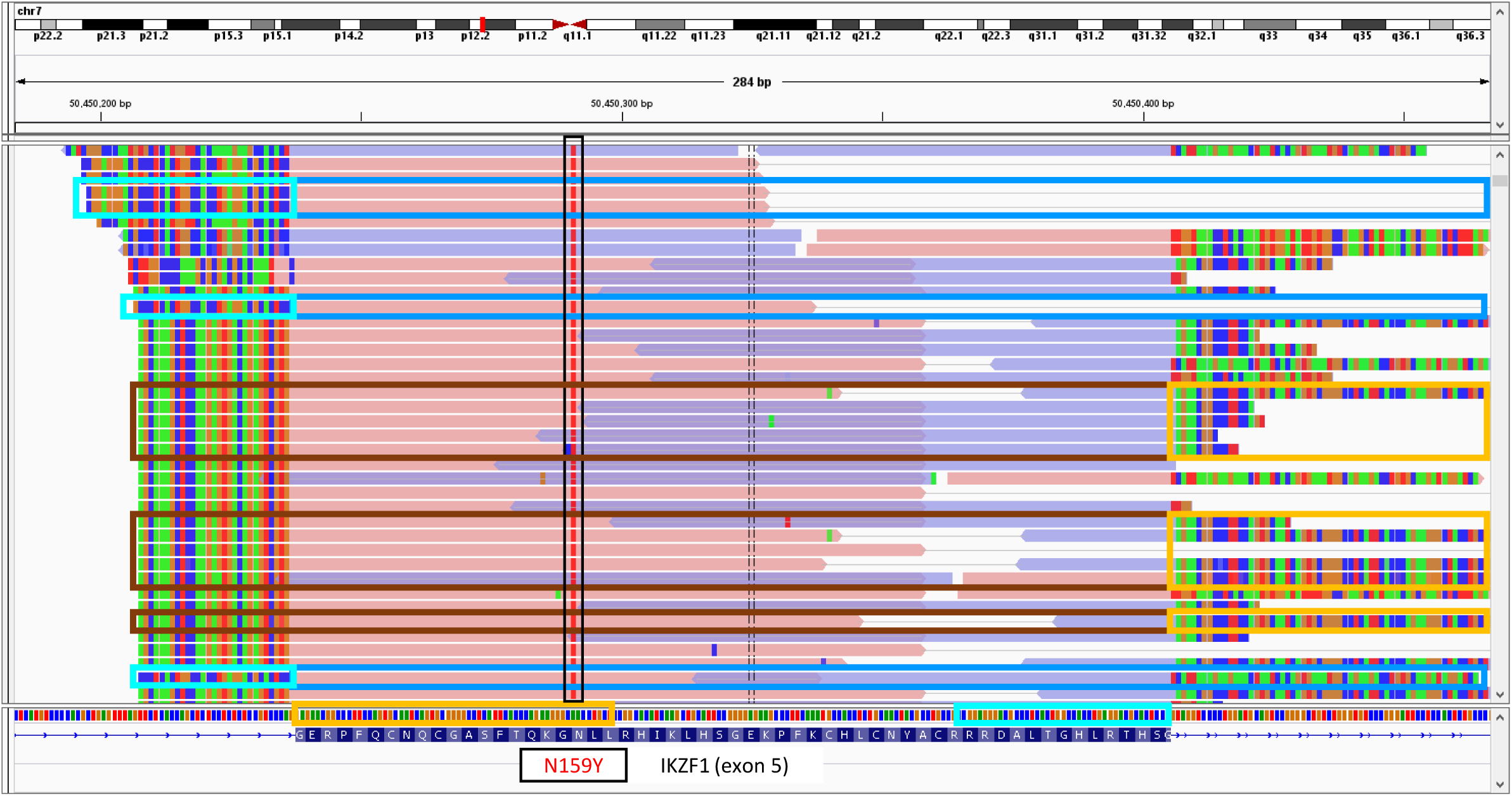
*IKZF1* e5e5 transcripts with duplication of N159Y. Both B-ALL cases with *IKZF1* N159Y (red positions in the reads, all surrounded by black box) demonstrated outlier expression of e5e5 where N159Y was present both 5’ of the e5e5 junction (reads surrounded by dark brown boxes) and 3’ of the e5e5 junction (reads surrounded by dodger blue boxes). Note that the softclipped ends of the reads (yellow and cyan boxes) match the 5’ and 3’ ends of exon 5 respectively, consistent with e5e5 junctions.

**SFig 4.**
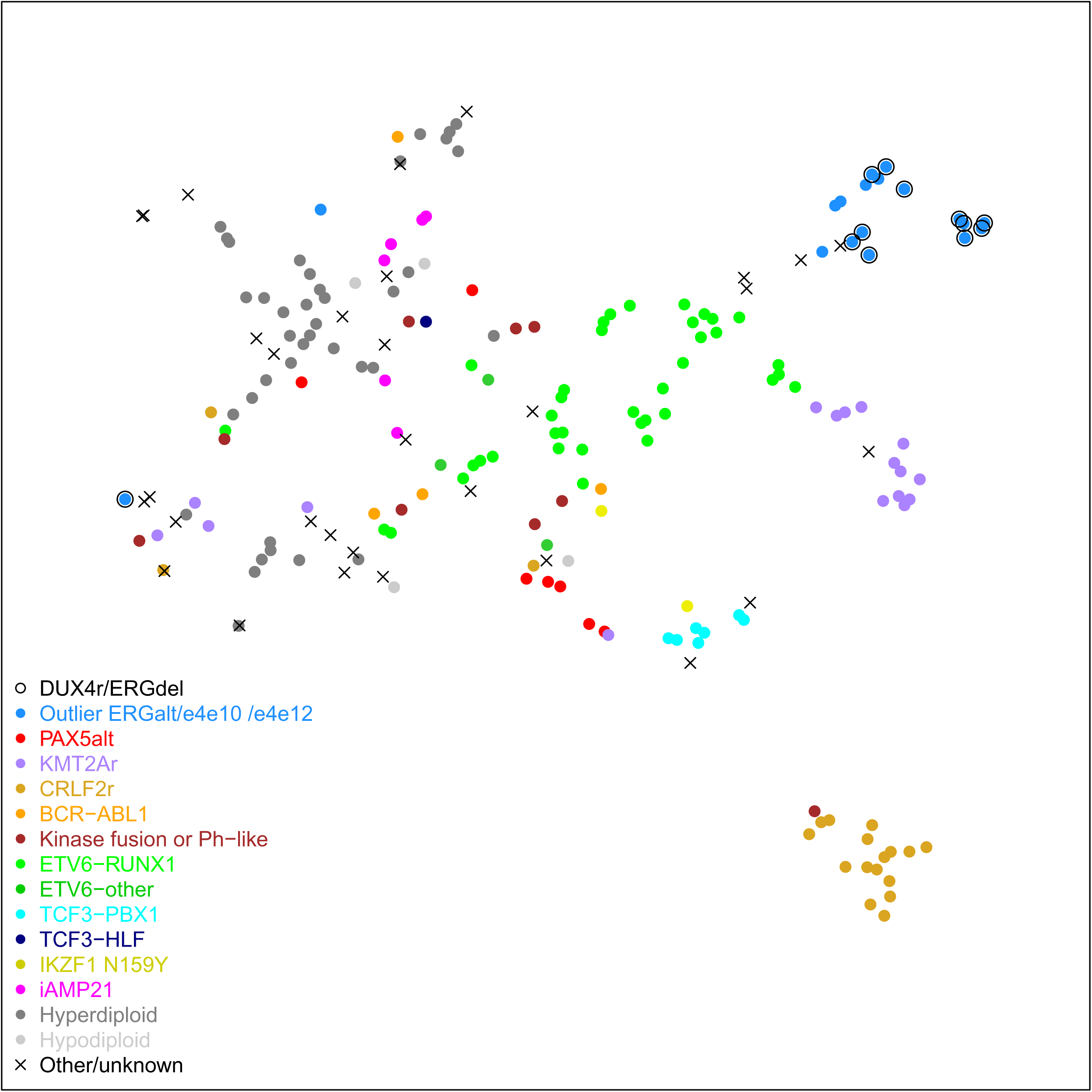
t-SNE of B-ALL cases relative to FPH expression of targeted genes. Under t-SNE applied to FPH gene expression data derived from the limited set of targeted genes, 11/12 B-ALL cases with known *DUX4* rearrangements and/or *ERG* deletions (dodger blue dots surrounded by open black circles) clustered together with 5/6 cases with outlier ERGaltA, e4e10, and/or e4e12 expression but unknown *DUX4*r status (dodger blue dots without surrounding circle). Two cases without outlier expression (black Xs) clustered nearby and did not have a well-defined subtype by molecular or cytogenetic studies; *DUX4*r status was negative by wtRNAseq in 1 of the cases and unknown in the other case. The remaining known *DUX4*r case (separate dodger blue dot surrounded by open circle) had lower cellularity and borderline RNA quality, and clustered nearby other cases with low blast percentage and/or borderline RNA quality. The final case with outlier ERGaltA expression (separate dodger blue dot without surrounding circle) had the DUX4/ERG signature by LDA card but was notable for its relapse status, suggesting the possibility of altered expression from progression events. Similar overall clustering patterns were observed under hierarchical clustering but were considerably less apparent by PCA or UMAP.

**SFig 5.**
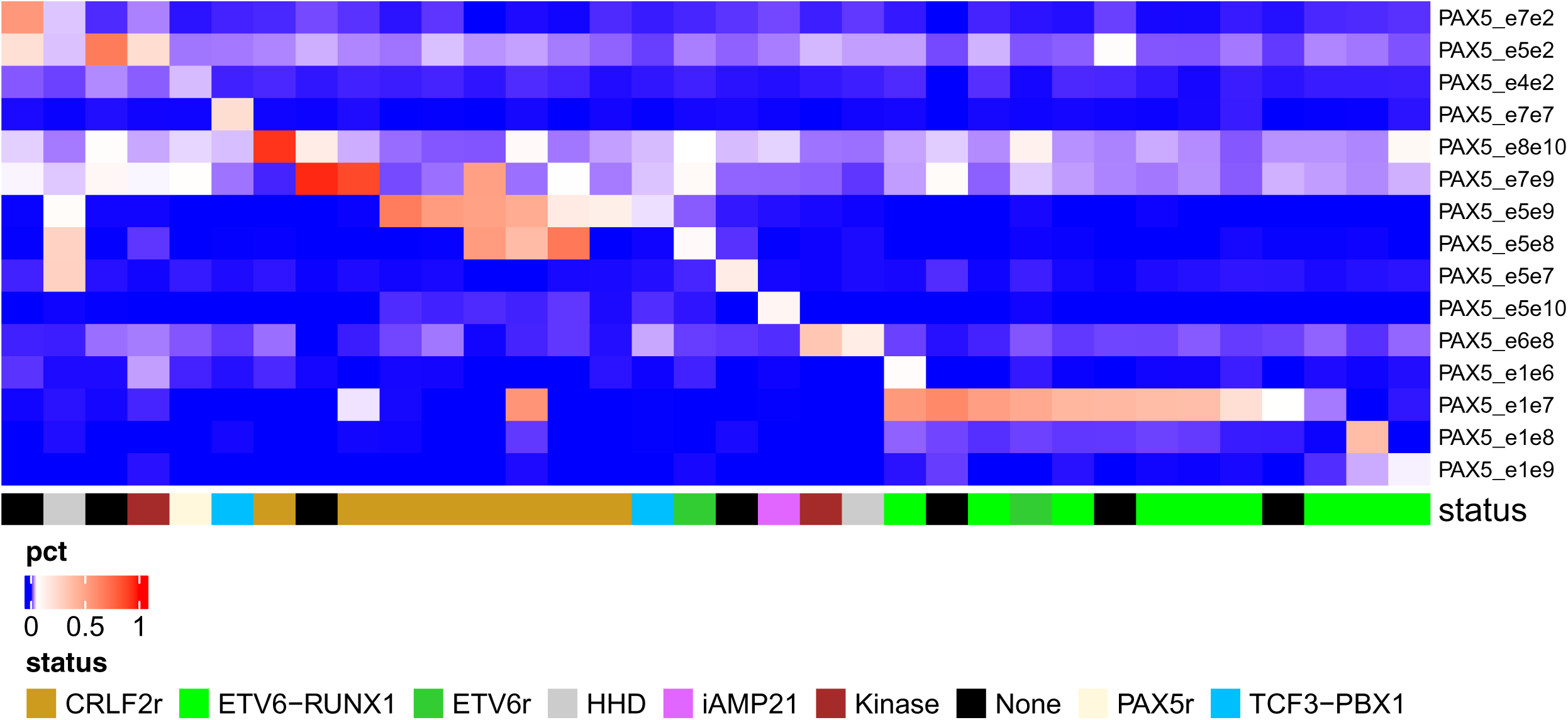
Fusions associated with outlier expression of *PAX5* isoforms. Cases with the highest VAFs (> 50%) of *PAX5* e5e2 or e7e2 were not associated with fusions or cytogenetically defined subtypes, favoring PAX5amp, whereas cases with moderate VAFs (~20%) of e5e2 or e7e7 were associated with a *PDGFRB* fusion and *TCF3-PBX1* respectively. Outlier expression of e5e8, e5e9, e7e9, or e8e10 was associated with *CRLF2* rearrangements while outlier expression of PAX5 e1e6, e1e7, e1e8, or e1e9 was associated with *ETV6-RUNX1*.

**SFig 6.**
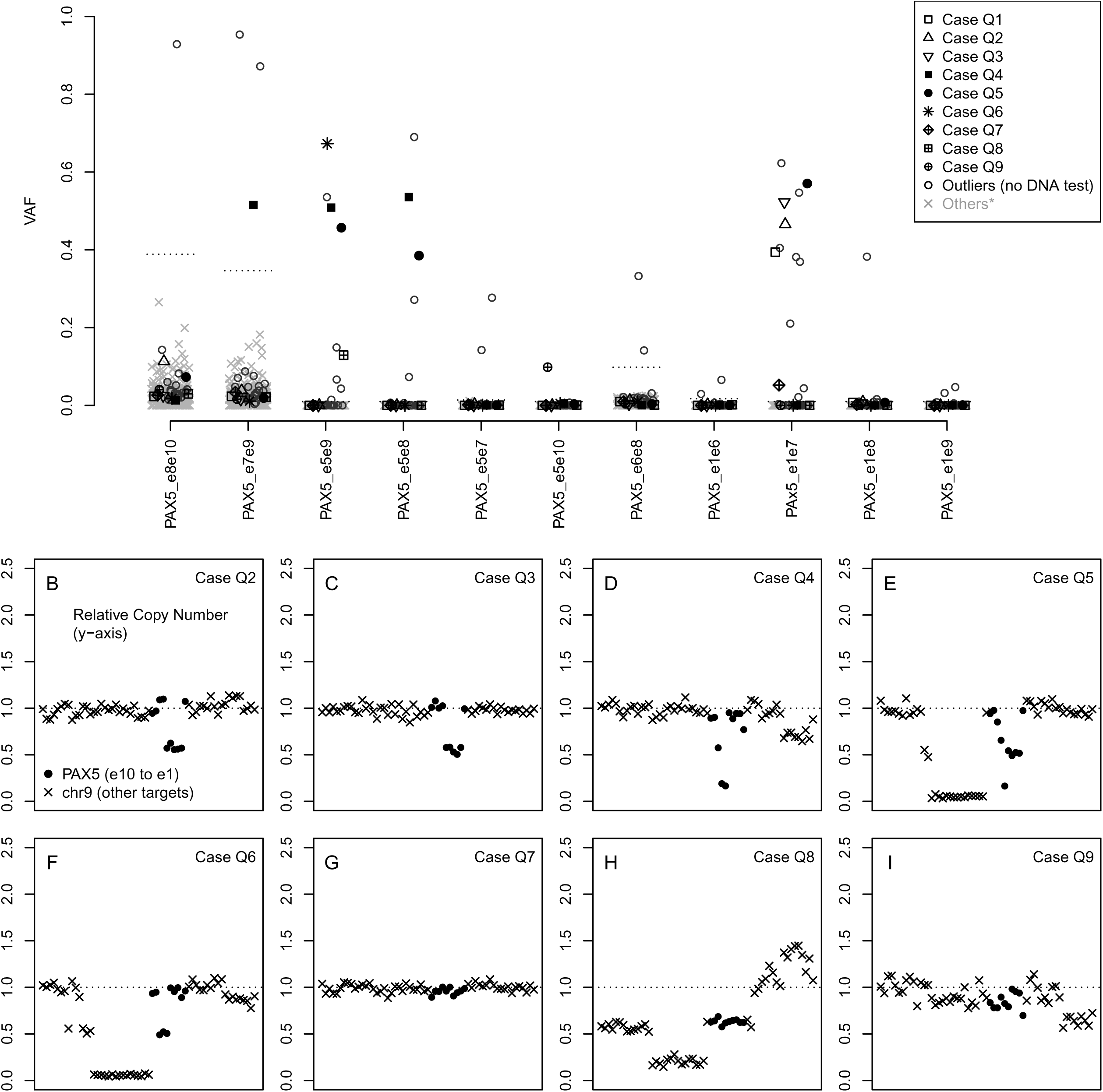
*PAX5* exon-skipping isoforms are associated with intragenic deletions. (A) Outlier expression of *PAX5* exon-skipping isoforms. Outlier thresholds were relatively high (close to 40%) for junctions associated with annotated alternative isoforms (e7e9, e8e10), however high VAFs indicative of deletion and LOH were robustly identified. Other thresholds were usually at 1% VAF except for *PAX5* e6e8. Two cases (Q4 and Q5) had outlier expression of 3 isoforms at similar VAFs around 40%-60%. (B-I) Copy number losses by MLPA. Cases Q2 (B) and Q3 (C) had e1e7 VAFs near 50% and corresponding 1-copy deletions of exons 2-6 in ~100% of cells. Cases Q4 (D) and Q5 (E) with multiple isoforms accordingly demonstrated multiple copy number levels consistent with multiple loss events. Cases Q7-Q9 (G-I) had low outlier VAFs (<20%) and were discordantly negative for intragenic deletions by MLPA, however the limit of detection by MLPA has been previously described as 15-30%.

